# Soluble DNA Concentration in the Perfusate is a Predictor of Post-Transplant Renal Function in Hypothermic Perfused Kidney Allografts

**DOI:** 10.1101/2023.05.16.23289983

**Authors:** Sergio Duarte, Anne-Marie Carpenter, Matthew Willman, Duncan Lewis, Curtis Warren, Isabella Angeli-Pahim, Werviston De Faria, Georgios Vrakas, Ashraf El Hinnawi, Thiago Beduschi, Narendra Battula, Ali Zarrinpar

**Author notes:** Corresponding author: Ali Zarrinpar, 1329 SW 16th St, Gainesville, FL 32608. Conflicts of interest: none.

## Abstract

**Introduction:** Hypothermic machine perfusion (HMP) has greatly facilitated kidney allograft preservation. However, tissue damage still occurs during HMP, deleteriously affecting post-transplant graft function. Therefore, improved methods to assess organ quality and to predict post-transplant graft function and survival are needed. We propose that soluble DNA (sDNA) measured in HMP perfusate can used as a non-invasive biomarker for this purpose.

**Methods:** Perfusate samples of kidney grafts placed on HMP were collected after 5 minutes and at the conclusion of HMP. sDNA of nuclear origin within the perfusate was quantified by real-time polymerase chain reaction and correlated with HMP parameters and post-transplant clinical outcomes.

**Results:** Kidney grafts from 52 donors placed on HMP were studied. Perfusate sDNA concentration was significantly elevated in transplanted kidneys with delayed graft function. Grafts with higher concentrations of perfusate sDNA at 5min and at HMP conclusion also had reduced graft function in the initial post-transplant period, as measured by post-operative day 2, 3, and 4 creatinine reduction ratios (CRR). Standard pump parameters such as renal vascular resistance and renal vascular flow were poor indicators of early post-transplant graft function.

**Conclusion:** sDNA concentration in HMP perfusate of kidney grafts can predict the quality of kidney graft preservation and indicate post-transplant renal function. This biomarker should be explored further to improve renal organ assessment and transplantation outcomes.

## Introduction

Advances in allograft preservation within the last century have transformed the field of solid organ transplantation. In renal transplantation, the widespread use of hypothermic machine perfusion (HMP) has become standard of care in kidney preservation, maintaining organ viability in the transition from donor to recipient and greatly reducing postoperative delayed graft function in the recipient compared to static cold storage (SCS).^1–3^ Additionally, HMP allows for longer storage times compared to SCS,^4^ providing the opportunity to continuously assess intrinsic graft attributes and levels of tissue damage that contribute to poor graft function, which is associated with recipient morbidity and mortality.^5^ Allografts continue to accrue injury during the preservation period; as such the quality of the graft upon procurement will not be the same after 24 to 48 hours of preservation. Thus, demand is high for methods to assess graft quality repeatedly during the preservation period and to predict graft function after transplantation.

In renal allografts preserved with HMP, methods of predicting donor organ function have had marginal success in reliably predicting organ damage and subsequent graft function. Currently, the most widely used method relies on dynamic parameters measured by the perfusion machine, namely, vascular resistance and flow, where flow correlates positively with graft function and resistance correlates negatively.^6^ However, the use of these indices to guide acceptance or rejection of donated kidneys has been criticized, as many allografts that may have been discarded on the basis of unacceptable pump parameters later demonstrate sufficient function after transplantation.^7–9^

To aid decision-making regarding donor organ acceptance, new avenues are being explored. Some have adopted a composite score assessing macroscopic appearance, flow, and urine output, when kidney grafts are subject to normothermic perfusion.^10^ Additional studies have focused on measuring biomarkers in the HMP perfusate, such as lactate dehydrogenase, aspartate transaminase, glutathione-S-transferase, interleukin-18, and other surrogates for glucose metabolism, oxygen consumption, glycolytic activity, ATP depletion, and mitochondrial damage.^11–13^ However, none of these dynamic values or biomarkers have demonstrated adequate power to predict post-transplant kidney function.^14, 15^ To date, a reliable method to directly measure renal allograft injury and predict postoperative renal function has not been established.

While no biomarker of machine-perfused donor organ injury and subsequent recipient allograft function has been validated for kidneys, some success has been achieved in lung transplantation. In a recent study, Kanou et al. measured the amount of cell-free DNA (herein referred to as soluble DNA or sDNA to avoid confusion with donor-derived cell-free DNA being quantified in recipient blood) in the perfusate of human donor lungs undergoing *ex vivo* HMP; they found that the concentration of sDNA was significantly higher in perfusates from lungs that developed severe primary graft dysfunction.^16^ These findings are echoed in a pre-clinical renal allograft study performed by our group. We quantified perfusate sDNA within porcine kidneys undergoing HMP and found that the amount of sDNA measured was directly proportional to histologic features of tissue necrosis.^17^ These findings served as the foundation for the study described herein.

In this study, we posit that sDNA measured within the HMP perfusate of human kidney allografts can be used as a non-invasive biomarker for organ integrity and as a result is able to predict post-transplant renal function.

## Methods

### Study Design

This is a single-center, prospective cohort study approved by the Institutional Review Board at the University of Florida (IRB#202001674). All human kidneys preserved by HMP as standard-of-care between July 2021 and December 2022 were included. Kidneys intended for pediatric (<18 years) recipients, discarded kidneys that were not transplanted, kidneys that were pumped at another institution prior to arriving at our institution, and kidneys involved in multi-organ transplants were excluded. Fifty-two kidneys and recipients were included in this study.

HMP parameters measured and registered for this study were renal vascular resistance (RR) and flow (RF) at initiation, 2 hours, 4 hours (if applicable), and endpoint of perfusion. The primary endpoint of the study was delayed graft function (DGF), defined as the need for dialysis within the first 7 days after transplantation. Secondary endpoints were post-transplant clinical outcomes indicative of early graft function such as estimated glomerular filtration rate (eGFR), creatinine (Cr), and creatinine reduction ratio (CRR, defined in Equation 1), which were measured on postoperative days (POD) 1, 2, 3, and 4. We restricted our analysis of early graft function to the first 4 postoperative days at the end of which most of the study subjects are discharged at our institution; the clinical outcomes datasets and study N decreases for each subsequent time point. A schematic of the study methodology is depicted in Figure 1.

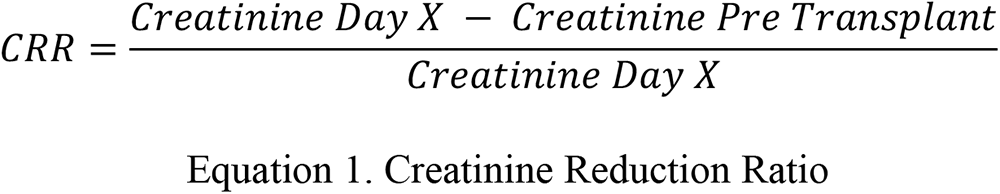

**Figure 1.**
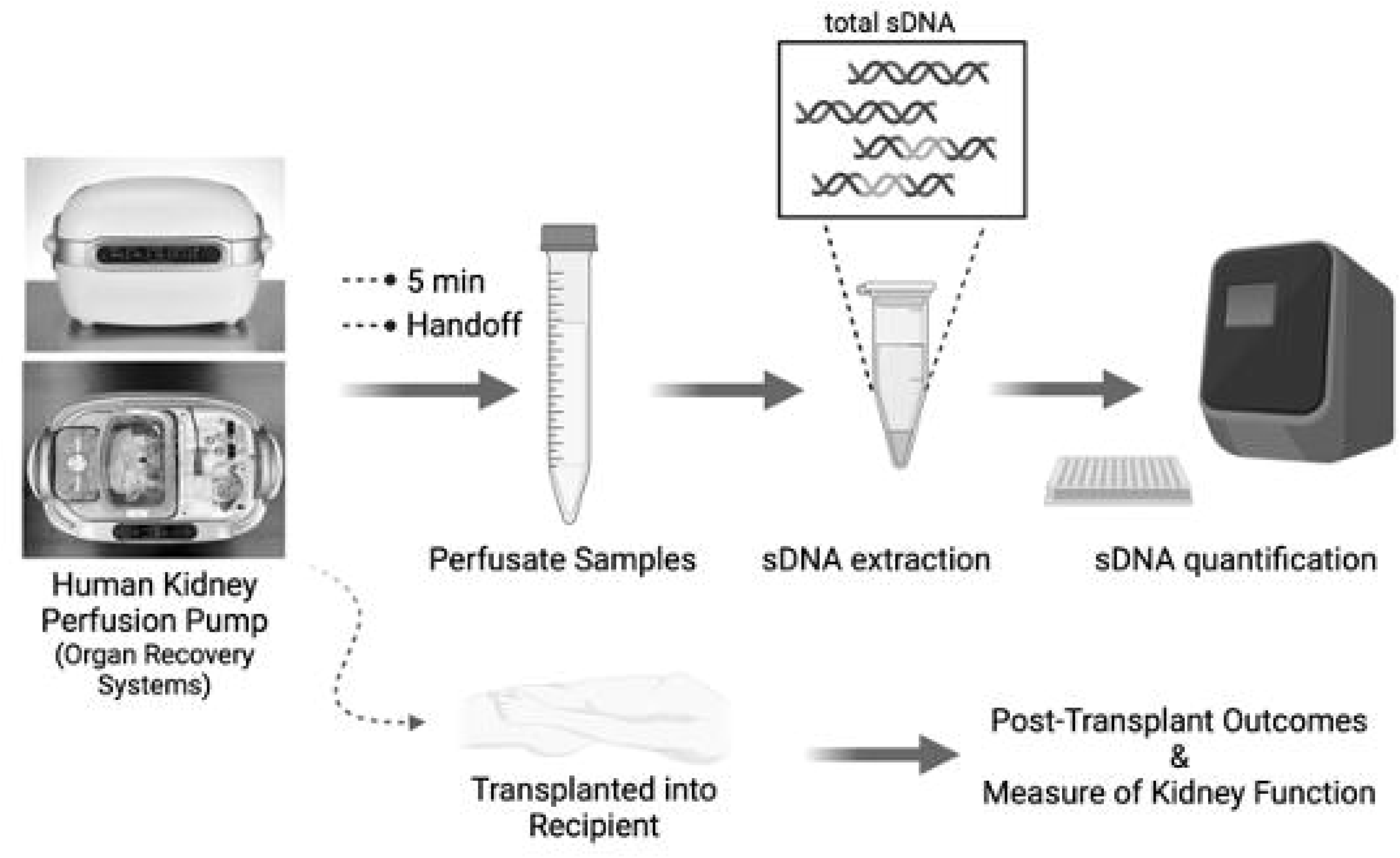
Study Methods. The perfusate of human kidneys undergoing hypothermic machine perfusion was sampled, and soluble DNA (sDNA) was extracted and quantified. After the kidney was transplanted into the recipient, post-transplant outcomes and measures of kidney function were assessed.

### Clinical Ex vivo Kidney Machine Perfusion Procedure

Kidneys utilized during the study period underwent HMP using the LifePort Kidney Transporter 1.1 (Organ Recovery Systems, Itasca, IL), according to manufacturer instructions and as described in previous investigations.^18, 19^ UW Machine Perfusion solution was used as perfusate. Perfusate samples of the kidney grafts selected for HMP were collected after 5 minutes of perfusion and at the conclusion of HMP at graft handoff to the surgical team for implantation.

### sDNA Extraction and PCR Quantification

The sDNA within each HMP perfusate sample was isolated using the QIAamp MinElute ccfDNA Mini Kit (QIAgen Group, Germantown, MD) according to manufacturer instructions. Briefly, 2mL perfusate was added to the proprietary magnetic bead suspension, which allows for binding of cell-free DNA to magnetic beads. The bound cell-free DNA was then eluted from the beads and purified using the QIAamp MinElute membrane. Purified cell-free DNA eluted from the membrane is the resulting soluble DNA (sDNA) sample. The nuclear-origin sDNA within the eluate was then quantified by real-time polymerase chain reaction (RT-PCR) using specific primer sequences for DNA of nuclear origin (customized oligonucleotide targeting GAPDH gene, Integrated DNA Technologies, Coralville, Iowa).

### Statistical Analysis

Statistical analysis was performed using the R software package (V.4.1.3, The R Foundation for Statistical Computing). Non-parametric Spearman correlations were used to assess the relationships (direction and strength) between sDNA concentration and HMP parameters, as well as between sDNA concentration and postoperative variables. Linear regression analysis was used to assess the effect of sDNA concentration on these outcomes.

## Results

### Graft, Donor and Recipient characteristics

A total of 52 kidneys and 52 recipients were studied. Demographic characteristics of donors and recipients are reported in Table 1. The majority of donor allografts were obtained from male (60.8%) donors after brain death (75.5%). Donor kidneys had a mean 640±309 min of static preservation on ice prior to placement on HMP, a mean 477±256 min of HMP, and a mean 1127±405 min of total cold ischemia. There was a significant relationship only between perfusate sDNA concentration at HMP conclusion (hereafter referred to as handoff) and total cold ischemia time (Table 2; ρ=0.3049). There were five cases of DGF among the recipients within the study period.

**Table 1.**
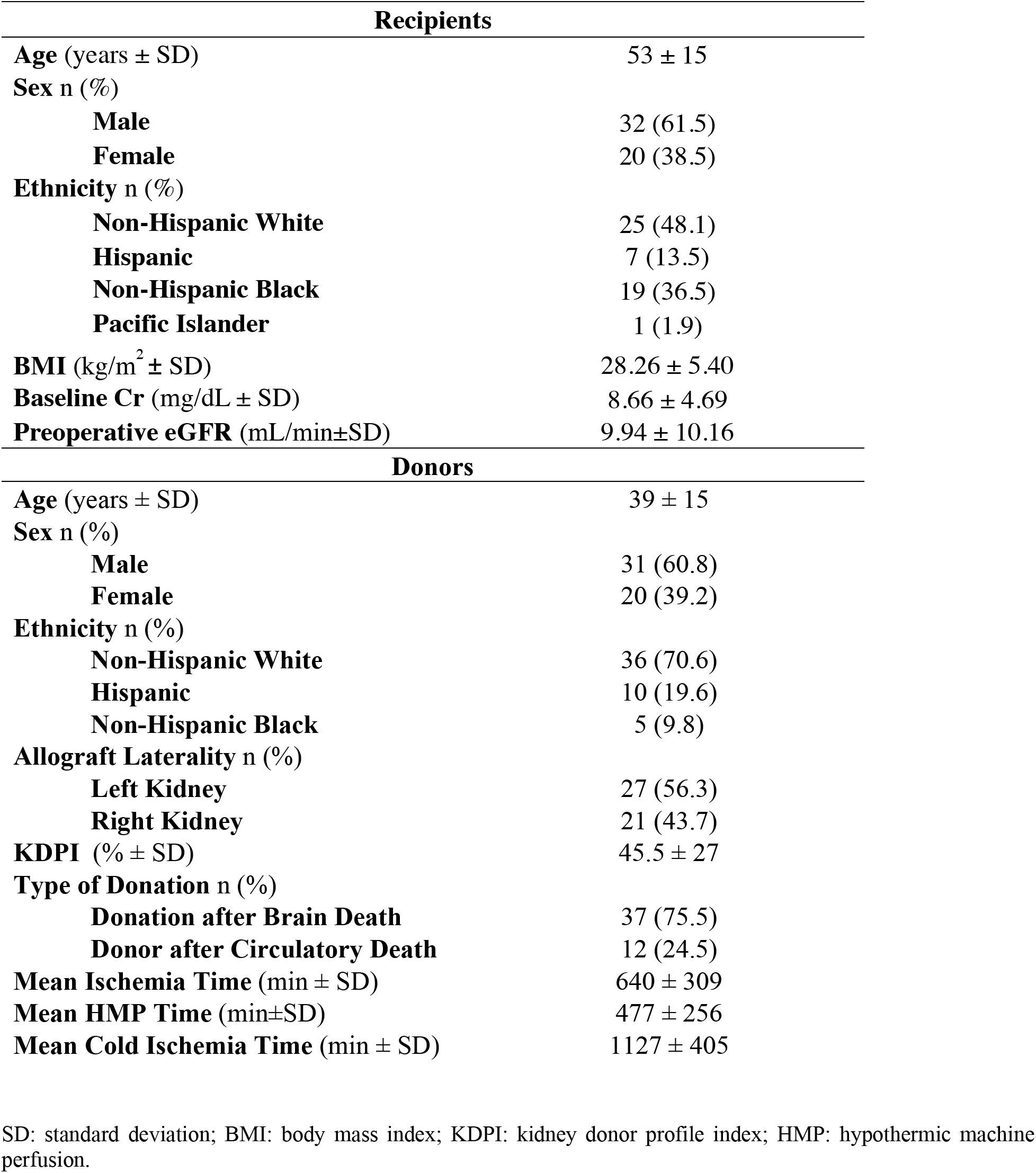
Donor and recipient demographics. Some donor kidney totals (n) do not add up to the total number of transplants performed (n=52), as this information was absent from the kidney donor information sheet at the time of data collection.

**Table 2.** sDNA levels and ex vivo organ timepoints.

| sDNA Concentration (ng/ml) vs Ischemic Parameters |  |  |  |
| --- | --- | --- | --- |
| Timepoint | Variable | Spearman Correlation ($\rho$) | p-value |
| **5 min** | Static Ice Time | 0.0287 | 0.847 |
|  | HMPT | 0.149 | 0.313 |
|  | CIT | 0.1486 | 0.313 |
| **Handoff** | Static Ice Time | 0.0877 | 0.5404 |
|  | HMPT | 0.238 | 0.0928 |
|  | CIT | **0.3049** | **0.0296** |

HMPT: hypothermic machine perfusion time; CIT: cold ischemia time (CIT).

### Association between ex vivo perfusion variables and post-operative graft function

In keeping with previous findings that standard means of measuring HMP donor organ integrity are poor predictors of post-transplant clinical measure of graft function, Table 3 presents the association of these variables with early postoperative Cr, eGFR, and CRR. There was no significant relationship between RF and any clinical measure of early post-transplant graft function. In contrast, we did observe a significant correlation between endpoint RR and Cr on all postoperative days up to day 4. Furthermore, neither RF or RR correlated significantly with CRR on any of the assessed postoperative days.

**Table 3.** Comparing post-transplant outcomes with renal flow, renal resistance, and kidney donor profile index.

| Endpoint Renal Flow |  |  |  |
| --- | --- | --- | --- |
| Timepoint | Outcome Variable | Spearman Correlation ($\rho$) | p-value |
| POD 1 | Cr | -0.166 | 0.234 |
|  | eGFR | 0.212 | 0.128 |
|  | CRR | -0.157 | 0.263 |
| POD 2 | Cr | -0.097 | 0.486 |
|  | eGFR | 0.091 | 0.515 |
|  | CRR | -0.119 | 0.398 |
| POD 3 | Cr | -0.020 | 0.890 |
|  | eGFR | 0.007 | 0.959 |
|  | CRR | -0.181 | 0.199 |
| POD 4 | Cr | -0.137 | 0.393 |
|  | eGFR | 0.210 | 0.183 |
|  | CRR | -0.106 | 0.511 |
| Endpoint Renal Resistance |  |  |  |
| Timepoint | Outcome Variable | Spearman Correlation ($\rho$) | p-value |
| POD 1 | Cr | **0.309** | **0.027** |
|  | eGFR | -0.256 | 0.069 |
|  | CRR | 0.091 | 0.525 |
| POD 2 | Cr | **0.302** | **0.031** |
|  | eGFR | **-0.293** | **0.037** |
|  | CRR | 0.075 | 0.598 |
| POD 3 | Cr | 0.216 | 0.140 |
|  | eGFR | -0.233 | 0.099 |
|  | CRR | 0.112 | 0.441 |
| POD 4 | Cr | 0.122 | 0.455 |
|  | eGFR | -0.182 | 0.260 |
|  | CRR | 0.229 | 0.161 |
| KDPI (%) |  |  |  |
| Timepoint | Outcome Variable | Spearman Correlation ($\rho$) | p-value |
| POD 1 | Cr | **0.307** | **0.028** |
|  | eGFR | **-0.286** | **0.042** |
|  | CRR | -0.160 | 0.262 |
| POD 2 | Cr | **0.379** | **0.0061** |
|  | eGFR | **-0.394** | **0.0042** |
|  | CRR | -0.198 | 0.164 |
| POD 3 | Cr | **0.377** | **0.0069** |
|  | eGFR | **-0.392** | **0.0045** |
|  | CRR | -0.161 | 0.265 |
| POD 4 | Cr | **0.418** | **0.0082** |
|  | eGFR | **-0.395** | **0.012** |
|  | CRR | -0.145 | 0.377 |

HMP: hypothermic machine perfusion; POD: postoperative day; KDPI: kidney donor profile index; Cr: Creatinine; eGFR: estimated glomerular filtration rate; CRR: creatinine reduction ratio.

### Association between KDPI and post-operative graft function

The Kidney Donor Profile Index (KDPI) is a cumulative percentile measure that characterizes the donor associated risk of post-transplant graft failure and aids transplant physicians in their decision to transplant a graft.^20^ However, the impact of HMP on KDPI’s association with early graft function is still unknown. When the KDPI was examined, we observed statistically significant correlations between KDPI and Cr, as well as eGFR on all postoperative days (Table 3). In contrast, there was no notable statistically significant relationship between KDPI and CRR as a measure of early post-operative graft function.

### Perfusion parameters correlate with perfusate sDNA

Next, we investigated whether standard perfusion measures that are currently used to assess organ quality correlated with perfusate sDNA levels. To do so, we compared sDNA at five minutes and at handoff to RR and RF at the start of HMP, at 2-hour and 4-hour time points, and at the conclusion of HMP (Supplementary Table 1). Five-minute (5min) perfusate sDNA levels correlated positively with graft RF on the pump at 2 hours and 4 hours, while handoff sDNA correlated with 2-hour, 4-hour, and final RF (Supplementary Table 1, Supplementary Figure 1). There was a negative trend in the correlations between RR and sDNA at 5min and at handoff, although these were not significant.

### Perfusate sDNA levels correlate with measures of early graft function

We then turned our attention to assessing whether perfusate sDNA levels correlate with post-transplant function. There was a statistically significant correlation between 5min sDNA and post-transplant graft function, with higher 5min perfusate sDNA concentrations correlating with lower CRR on POD2 and POD4 (Supplementary Table 2, Supplementary Figure 2).

Importantly, grafts with higher concentration of perfusate sDNA at time of graft handoff had significantly reduced early post-transplant graft function (Table 4, Figure 2). Specifically, higher handoff sDNA concentrations correlated strongly with lower CRR on all postoperative days, lower POD3 eGFR, and higher POD3 and POD4 serum Cr levels.

**Figure 2.**
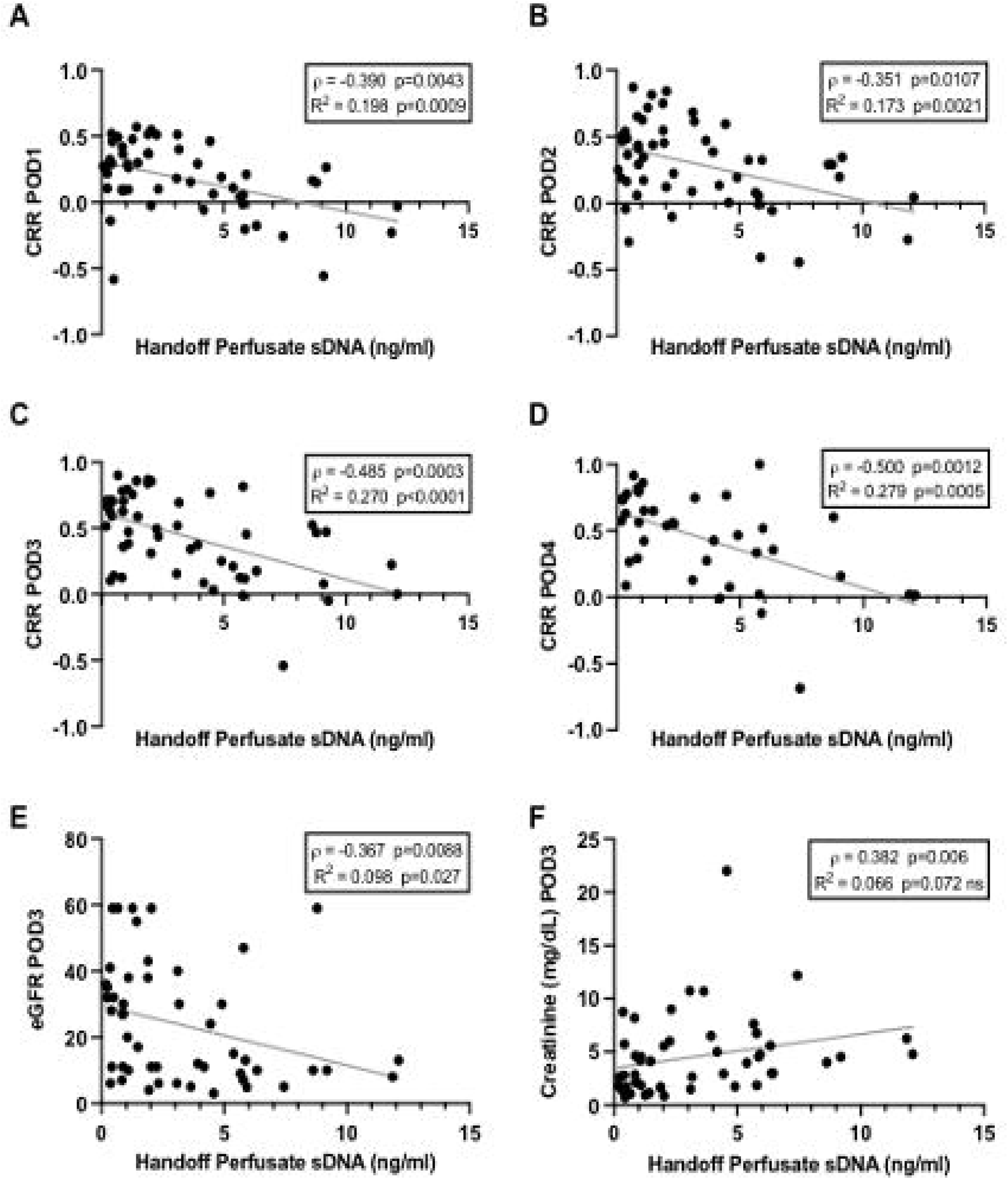
Handoff sDNA vs measurements of early graft function. Handoff sDNA negatively correlated with creatinine reduction ratio (CRR) on all postoperative days (POD) (A-D). Handoff sDNA showed a similar negative correlation with estimated glomerular filtration rate (eGFR) on POD3 (E) and POD4 (not shown), as well as a positive correlation with POD3 (F) and POD4 Creatinine.

**Table 4.** Handoff sDNA concentration correlation with early graft function.

| Timepoint | Outcome Variable | Spearman Correlation ($\rho$) | p-value |
| --- | --- | --- | --- |
| POD 1 | Cr | 0.041 | 0.774 |
|  | eGFR | -0.107 | 0.457 |
|  | CRR | **-0.390** | **0.0043** |
| POD 2 | Cr | 0.214 | 0.131 |
|  | eGFR | -0.256 | 0.069 |
|  | CRR | **-0.351** | **0.0107** |
| POD 3 | Cr | **0.382** | **0.006** |
|  | eGFR | **-0.367** | **0.0088** |
|  | CRR | **-0.485** | **0.0003** |
| POD 4 | Cr | **0.330** | **0.043** |
|  | eGFR | **-0.373** | **0.019** |
|  | CRR | **-0.500** | **0.0012** |

POD: postoperative day; Cr: creatinine; eGFR: estimated glomerular filtration rate; CRR: creatinine reduction ratio.

### sDNA levels predict delayed graft function

Finally, we examined the relationship between sDNA in HMP perfusate and the development of DGF postoperatively. There was a significantly higher level of handoff sDNA (p=0.018) and 5min sDNA (p=0.047) in the kidneys whose recipients ultimately exhibited DGF in comparison with the kidneys that did not exhibit DGF (Figure 3A, 3B). The receiver operating characteristics (ROC) curve for our cohort determined that sDNA concentration as a biomarker for DGF has a reliable prognostic performance with an area under the curve (AUC) of 0.816 (95% CI 0.68-0.96; p=0.021) in samples obtained at kidney handoff to surgeon, and an AUC of 0.771 (95% CI 0.58-0.96; p=0.048) in samples obtained at 5min of perfusion. Based on a Youden index/ROC curve analysis of the cohort, using 2.69 ng/ml of sDNA in the handoff perfusate as the threshold for the likelihood of post-transplant DGF yields 100% sensitivity (95% CI 0.57-1.00) and 64.7% specificity (95% CI 0.51-0.76) (Figure 3C, 3D). In comparison, the ROC curves generated for KDPI, RR, and RF in predicting DGF were less reliable and were not statistically significant with this cohort (Figure 3E, 3F, 3G).

**Figure 3.**
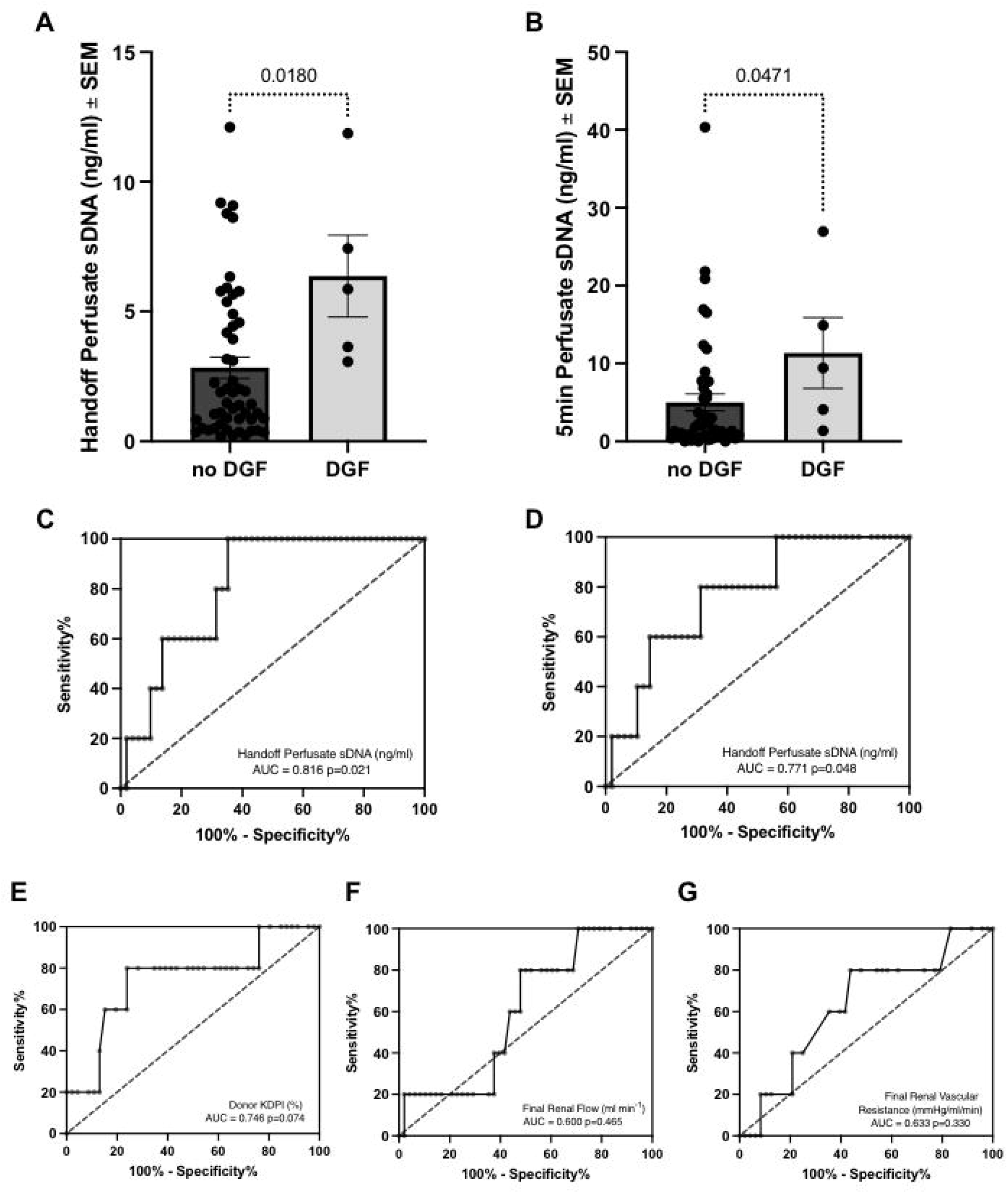
Soluble DNA levels in predicting delayed graft function (DGF). Handoff (A) and 5min (B) sDNA concentration was significantly higher in the five individuals who exhibited DGF. The receiver operating characteristic (ROC) curves for our cohort demonstrate that sDNA as a biomarker for DGF has a reliable prognostic performance with an area under curve (AUC) of 0.816 for samples obtained at handoff (C) and AUC=0.771 for samples obtained at 5min perfusion (D). Conversely, the ROC curves for Kidney Donor Profile Index (KDPI, E), renal vascular flow (F), and renal vascular resistance (G) were less reliable and were not statistically significant (KDPI AUC=0.748, p=0.074; flow AUC=0.600, p=0.485; resistance AUC=0.633, p=0.330)

## Discussion

The utilization of kidney allografts is left to the discretion of transplant physicians. This can be a daunting task, as the recipient’s health and quality of life is significantly affected by the outcome of this decision.^21^ Several factors, such as KDPI, appearance, ischemia time, histologic features of any biopsy, and dynamic values such as RR and RF produced during HMP, are used to inform the decision. However, there is no quantitative, noninvasive, repeatable biomarker of tissue damage correlating with postoperative renal function available to aid in the decision-making process. In this study, we propose sDNA measured in HMP perfusate of renal allografts as one such biomarker.

Soluble DNAs are cell-free, circulating, short fragments of DNA released from injured, necrotic, apoptotic, and other dying cells. Soluble DNA can be measured and characterized in plasma and is revolutionizing many medical fields such as oncology, maternal fetal medicine, and transplantation.^22–24^ The measurement of DNA has become a useful practice in determining allograft integrity in the post-transplant setting.^25^ Several studies have validated that donor-derived cell-free DNA can be quantified in the bloodstream of renal transplant recipients and used as a surrogate for graft injury.^26–28^ As it has been shown to be a biomarker for graft damage in the recipient bloodstream, it is likely that measurement of soluble DNA in the perfusate during HMP could also provide insight as to graft damage prior to transplantation. This was very recently demonstrated in pulmonary allografts,^16^ but has not been shown within human donor kidneys for which HMP is widely established as a clinical standard, until now.

Here, we confirm that sDNA is a reliable measure of early graft function in a post-transplant population. We demonstrate that sDNA within the HMP perfusate correlates with markers of renal function post-transplant. Specifically, higher 5-minute sDNA concentrations correspond with lower CRR on POD2 and POD4 (Supplementary Table 2, Supplementary Figure 2), and a higher level of sDNA at handoff significantly correlates with a lower CRR on all postoperative days (Table 4, Fig 2). The rate of creatinine clearance after transplantation is a more relevant assessment of short-term graft function than simple creatinine levels. CRR is an accurate of measure of this rate. Additionally, CRR calculated on POD2 has previously been shown to predict long-term graft outcomes, specifically serum Cr at one year and at 5 years post-transplant.^29–31^ In comparison, none of the commonly employed predictors of a graft’s suitability (KDPI, RR, and RF) were significantly correlated with CRR on any postoperative day in this study (Table 3). We do note that higher KDPI was significantly correlated with lower eGFR in our study (Table 3), supporting its continued use to guide decisions regarding graft suitability in conjunction with other parameters such as sDNA concentration.

One finding within our study requires additional scrutiny, namely, the observation that renal flow is positively correlated with sDNA concentration, while renal resistance is negatively correlated (Supplementary Table 1, Supplementary Figure 1). While counterintuitive, one possible explanation for these results is that higher flow during HMP allows for greater exposure of damaged tissue to the HMP perfusate, allowing the nuclear-origin DNA to solubilize within the solution and thus be detected at higher concentration. This relationship between sDNA and pump parameters requires further investigation, but our findings nonetheless indicate that higher sDNA within HMP perfusate is associated with worse post-transplant outcomes, particularly lower eGFR and CRR (Supplementary Table 2, Supplementary Figure 2, Table 4, Figure 2) as well as higher rates of DGF (Fig 3), where RR and RF have previously proved unreliable.

Though exciting, the results of this study are limited by several factors, including its single-center nature and sample size. A larger, multi-center study would greatly improve demographic and practice diversity in addition to increasing the sample size. Furthermore, only 5 recipients demonstrated delayed graft function, limiting the clinical interpretation of our calculated specificity and sensitivity (Fig 3). Finally, though measures of early graft function were used as the primary outcomes, extension of the study period to longer than POD4 may reveal additional insights. Ongoing investigations will address these limitations. Despite these limitations, our findings indicate that sDNA is a promising predictor of post-transplant function in human renal allografts undergoing HMP. Future work will be devoted to incorporating these findings into clinical practice.

In conclusion, we demonstrate that the concentration of sDNA in the perfusate of *ex vivo* hypothermic perfused kidney grafts provides insight on the quality of these grafts at the time of transplantation. Furthermore, the sDNA levels are directly correlated with early post-transplant renal function. We propose that perfusate sDNA should be explored as a candidate biomarker for tissue damage and early graft function in hypothermic machine perfused renal allografts.

## Data Availability

All data produced in the present study are available upon reasonable request to the authors

## Disclosures

The authors have nothing to disclose.

## Funding

This study was supported by an ASTS-CareDx Career Development Grant and by NIH/NIDDK K08DK113244 (AZ).

## Notes

### Competing Interest Statement

The authors have declared no competing interest.

### Author Declarations

The Institutional Review Board of University of Florida gave ethical approval for this work

