## Supplemental tables and figures for "Soluble DNA Concentration in the Perfusate is a Predictor of Post-Transplant Renal Function in Hypothermic Perfused Kidney Allografts"

### Supplementary Information

#### Table of Contents

Supplementary Table 1……………………………………………………………………1

Supplementary Figure 1…………………………………………………………………...2

Supplementary Table 2……………………………………………………………………3

Supplementary Figure 2…………………………………………………………………...4

**Supplementary Table 1.** Perfusate sDNA vs hypothermic machine perfusion parameters.

| sDNA Concentration and HMP Parameters | | | |
|---|---|---|---|
| **Timepoint** | **Variable** | **Spearman Correlation (ρ)** | **p-value** |
| **5 min** | Initial RR | -0.145 | 0.320 |
| | 2hr RR | -0.272 | 0.074 |
| | 4hr RR | -0.135 | 0.413 |
| | Final RR | -0.115 | 0.441 |
| **Handoff** | Initial RR | 0.0063 | 0.965 |
| | 2hr RR | -0.200 | 0.178 |
| | 4hr RR | -0.109 | 0.498 |
| | Final RR | -0.186 | 0.199 |
| **5 min** | Initial RF | 0.215 | 0.143 |
| | 2hr RF | **0.334** | **0.023** |
| | 4hr RF | **0.311** | **0.051** |
| | Final RF | 0.277 | 0.057 |
| **Handoff** | Initial RF | 0.128 | 0.371 |
| | 2hr RF | **0.391** | **0.0055** |
| | 4hr RF | **0.365** | **0.017** |
| | Final RF | **0.364** | **0.0087** |

HMP: Hypothermic machine perfusion; RR: renal vascular resistance; RF: renal vascular flow; 5min: concentration of sDNA at 5minutes hypothermic machine perfusion; Handoff: concentration of sDNA at endpoint hypothermic machine perfusion.

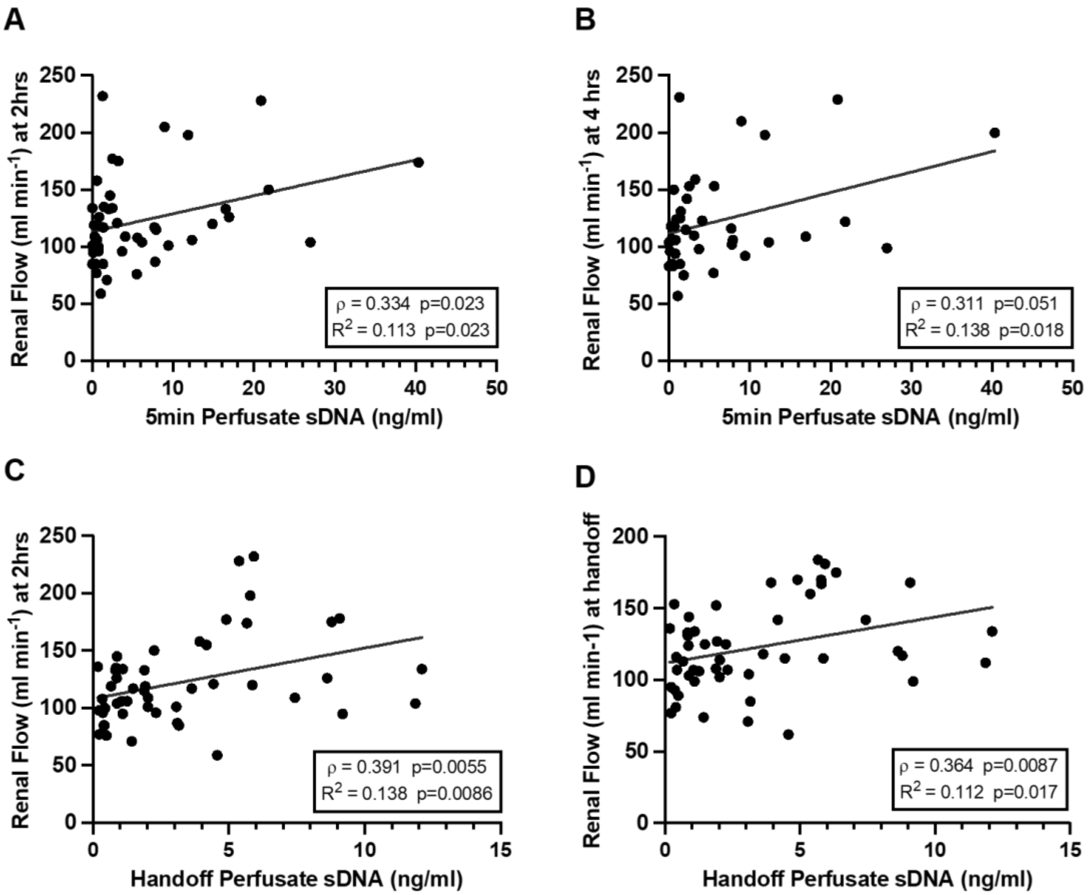


**Supplementary Figure 1. Graphical representation of dynamic hypothermic machine perfusion (HMP) parameters vs sDNA levels in the perfusate.** There was significant correlation between 5min sDNA with renal flow at 2hrs and at 4hrs (A, B), and between handoff sDNA and renal flow at 2hrs (C), 4hrs (not shown), and at handoff (D).

**Supplementary Table 2.** Five-minute sDNA concentration compared with early graft function.

| 5min sDNA Concentration and Early Graft Function | | | |
|---|---|---|---|
| **Timepoint** | **Variable vs 5min sDNA (ng/ml)** | **Spearman Correlation (ρ)** | **p-value** |
| **POD 1** | Cr | -0.049 | 0.743 |
| | eGFR | -0.066 | 0.646 |
| | CRR | -0.254 | 0.072 |
| **POD 2** | Cr | 0.121 | 0.413 |
| | eGFR | -0.127 | 0.389 |
| | CRR | **-0.314** | **0.025** |
| **POD 3** | Cr | 0.123 | 0.409 |
| | eGFR | -0.114 | 0.441 |
| | CRR | -0.267 | 0.061 |
| **POD 4** | Cr | 0.244 | 0.098 |
| | eGFR | -0.144 | 0.396 |
| | CRR | **-0.342** | **0.033** |

POD: postoperative day; Cr: creatinine; eGFR: estimated glomerular filtration rate; CRR: creatinine reduction ratio.

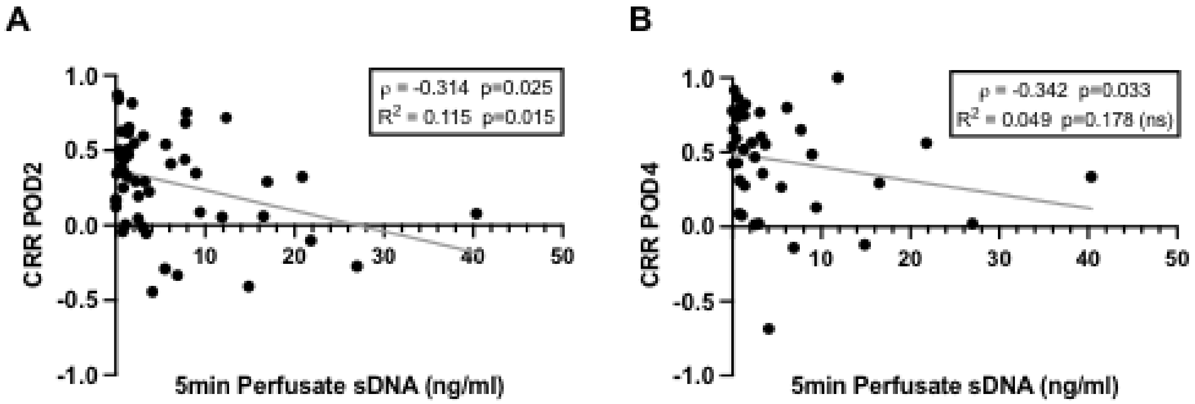


**Supplementary Figure 2. Five-minute sDNA concentration vs creatinine reduction ratio (CRR).** 5min sDNA negatively correlated with CRR on postoperative day (POD) 2 (A) and POD4 (B).
